# Older Age and Vaccination Protect Against Transaminase Elevation in Pediatric SARS-CoV2

**DOI:** 10.1101/2024.04.01.24303996

**Authors:** Antonia Fernandez Ovale, Cassandra Charles, Janet Rosenbaum, Priscila Villalba-Davila, Mauricio Mora, Shagun Sharma, Vivian Vega Lemus, Saema Khandakar, Thomas Wallach

## Abstract

**Objectives:** SARS-CoV2 infection is reported to induce transaminase elevations. There are case reports of severe liver injury in adult SARS-CoV2 patients and some have theorized that acute SARS-CoV2 infection may be a driver of severe liver injury in children. While pediatric hepatic injury has previously been described, clear shifts in immunogenic response secondary to prior immune exposure and vaccination since initial reports from 2020 warrant further evaluation. We sought to identify the impact of variant shifts and vaccination on this phenomenon in children.

**Methods:** A retrospective, cross-sectional study of pediatric SARS-CoV2 patients seen at two hospital facilities in an urban neighborhood in New York City between March 2020 and March 2022 was conducted via chart review. Data was extracted relating to patient’s demographics, clinical presentation, including the level of care and the laboratory results of comprehensive metabolic panels (CMP).

**Results:** 133 pediatric cases were identified as having positive SARS-CoV2 and CMP obtained in the same visit. Patients were predominantly Black (79.2%) and non-Hispanic (87%) with a mean age of 9.2 years. Risk of transaminase elevation was increased in younger patients and patients with higher level of care. BMI was not a risk factor noted for transaminase elevation. Vaccination decreased degree, not incidence, of transaminase elevation but given low rates of vaccination unable to determine significance of protective efficacy.

**Conclusions:** Our study has identified a profound increased risk of transaminase elevation in younger patients, the absence of BMI as a correlating factor in our primarily Black patient population, a shift towards non-specific AST elevation with variant windows and a strong signal of vaccine protection.

**What is Known:**

- SARS-CoV2 can cause Transaminemia, and in rare cases, possible fulminant hepatic injury
- Pediatric SARS-CoV2 infections are statistically milder than adult.
- SARS-Cov2 case severity and complications like multisystem inflammatory syndrome in children have declined over time

**What is New:**

- With population immunity and variant shifts transaminase elevations increasingly may not be of hepatic origin
- Risk of transaminase elevation is substantially higher in younger patients, and decreases with age
- Vaccines are protective against degree of transaminase elevation, and likely against incidence of transaminase elevation, although further study is needed.

**Article Summary:** Cross sectional study of an urban pediatric population demonstrates SARS-CoV2 transaminase elevation linked with younger age, unvaccinated status, and higher level of care.

## Introduction

It has been well-documented since the onset of the pandemic that SARS-CoV2 infections can trigger elevations to serum transaminases, thus raising concerns for direct liver injury^1^. While direct liver failure as a result of SARS-CoV2 appears to be rare and often perceived to be secondary to multi-organ injury in severely ill patients^2^, it is not fully elucidated how acute viral infection impacts the pediatric liver nor how this picture has changed over time with evolved variants and acquired population immunity^3^. In 2021, a perceived cluster of pediatric fulminant liver failure led to rampant speculation that SARS-CoV2 was driving incidence, and raising concern for further increase. Further investigation led to foundational work demonstrating the role of adeno-associated virus but no evidence of SARS-CoV2 involvement was determined besides seroprevalence, which at that time was known to be wide^4, 5^.

In the adult population, it has been reported that approximately 20 to 30% of patients who were hospitalized due to SARS-CoV2 acute infection presented with acute liver injury (ALI), characteristically presenting with mild cholestasis and elevation of transaminases^2^. While this rate may vary with disease severity and history of pre-existing conditions, it suggests that for adult patient’s liver involvement may be predictive of more severe disease^6^, although it is unclear whether the impact on the liver was due to either severe illness or viral infection directly. Mechanistic work has suggested that direct viral injury to the liver is plausible^7^. Pediatric case series studies have suggested the viruses have the potential to drive liver failure^8^ yet further understanding of the natural history of this disease in pediatrics is needed, especially as meaningful interventions such as vaccination have been deployed.

As rates of fulminant liver injury have remained essentially unchanged over time, it is unlikely that acute SARS-CoV2 infections have led to an increased need for liver transplantation or frequently driven direct morbidity and mortality from hepatic injury. However, it remains a serious possibility that SARS-CoV2 may present an ongoing risk for severe liver injury, especially given evidence of increased capacity to drive autoimmunity, including liver specific disease^9-11^. There is an absence of confirmed findings for relatively rare events such as pediatric liver failure that is not exculpatory of possible linkage. It is also likely that the pathology has shifted over time from when previous studies of viral interaction with the pediatric liver^12, 13^ were reported which is evidenced by overall decreasing clinical acuity and a rapid decline in Multisystem Inflammatory Syndrome in Children (MIS-C) with different waves of variant infection^14^.

In this study, we sought to retrospectively assess the natural history, population immunity and associated risks of acute lab-confirmed SARS-CoV2 infection on liver function at neighboring hospital facilities in an urban neighborhood in Brooklyn, NY between March 2020 and March 2022. As SARS-CoV2 vaccination rates remain low in children, our work is particularly relevant in understanding the value of vaccination in children which remains contentious and highly important^15^. We hypothesized that transaminase elevations would be associated with increased body mass index (BMI), a higher level of treatment care and more prevalent in the early phases of the epidemic. We anticipated milder symptomology and lower rates of transaminase elevation in those children improvement from prior immune exposure via infection or vaccination.

## Methods

SUNY Downstate IRB and the Kings County STAR Committee approved the retrospective review of the electronic medical records of pediatric patients evaluated for SARS-CoV2 at two neighboring New York City institutions between March 2020 and March 2022. Our review included all patients up to and including 18 years of age with confirmed SARS-CoV2 infection via nasal swab-derived real-time reverse polymerase chain reaction, as well as a comprehensive metabolic panel performed at the same visit. We reviewed over 3452 cases and identified 133 which met our criteria.

Clinical, demographic, laboratory and anthropometric data were collected. Patients were evaluated at the emergency department prior to being admitted to the pediatrics floor or pediatric intensive care unit. No patients were repeated. The following demographic covariates were extracted from the patient chart: age in months, male versus female sex assigned at birth, Black vs non-Black race, and Hispanic versus non-Hispanic ethnicity. We had the following health status measures: BMI, preexisting conditions, and asthma status.

Patients that were diagnosed with MIS-C during the hospitalization were excluded. Patients with sickle cell disease were also excluded from our analysis due to the possible increase of transaminases due underlaying hemolysis. All the patients included in our study that had a prior value of aspartate aminotransferase (AST) or alanine aminotransferase (ALT) available in electronic medical record, had normal values for transaminases prior to the acute infection by SARS-CoV-2. Patients with prior elevation of liver enzymes were excluded. AST and ALT elevation were assigned using cutoffs from Bussler et al (AST >46, ALT >20)^16^. Transaminemia trends were only followed up until discharge from the hospital. No follow up results were included once patients were discharged from the unit.

Patients were categorized into two groups, one with elevated transaminases at initial encounter or during hospitalization and another group that presented with normal transaminases at initial encounter and during the hospital course, if admitted.

Gastrointestinal symptoms were considered positive if the patient presented with at least one of the following: nausea, vomiting, diarrhea, abdominal pain and decrease oral intake. Obesity was defined as body mass index (BMI) above 95th percentile and/or BMI ≥30 kg/m^2^ as applicable. Level of care was determined depending on the admission location or discharged from the emergency department after initial encounter with no follow up.

SARS-CoV-2 variants were estimated variant type based on population sequencing data from New York State. This was estimated by grouping patients on timelines according to the first time each of the main variants was detected in New York until the time a subsequent variant was detected. Vaccination status of each patient was reviewed as per city wide immunization records of New York and the presence of SARS-CoV-2 antibody was accounted only when performed during the acute SARS-CoV-2 infection hospitalization.

### Analysis

We evaluated the association between elevated transaminases and continuous variables (age in months, AST/ALT ratio, BMI z-score) using Kolgomorov-Smirnov to evaluate the difference between the areas between the curves and the non-parametric Wilcoxon test. We used ggplot2 to produce plots, https://ggplot2.tidyverse.org) in R 4.3.1^17^. Elevated transaminases were present in 53/133 patients. Multivariate Poisson regression was used to assess the outcome of elevated transaminases in these patients.

Vaccination status is highly endogenous to age and date of presentation because the FDA issued emergency use authorization for the vaccine approvals by age groups, and the vaccine was first authorized for ages 12 and older on May 10, 2021 and for ages 5-11 on October 29, 2021, and the vaccine was not available for under 5 until June 2022. In our sample, only 3 patients ages 5-11 were vaccinated. We cannot distinguish between the effects of older age and vaccination status. SARS-CoV-2 antibody status was coded as true for children who tested positive on the antibody test and coded negative for children who either were not administered an antibody test or who tested negative on the antibody test: the test may have been differentially administered to children who had elevated transaminases.

We used causal mediation analysis to evaluate how much of the association between vaccination and elevated transaminases is mediated by age, using the mediation package^18^.

## Results

133 patients meeting our criteria were included. Patients age was from 0 to 18 years old at the time of encounter with a mean age of 9.2 years old. 52.9% of the patients were male and 45.5% of the patients did not have a comorbidity. Most of our study population was black and non-Hispanic with 79.2% and 87% respectively. No patients had progressive liver injury or evidence of liver failure or complications of liver injury.

56% of patients presented with one or more gastrointestinal symptoms. 47.2% patients with elevated transaminases presented with one or more gastrointestinal symptom at initial encounter. There was no association between the presence of gastrointestinal symptoms and transaminase elevation status (p-value = 0.20). There was also no statistical significance between vaccinated and non-vaccinated patients and the presence of gastrointestinal symptoms at onset (p-value = 0.2). There was no difference in BMI between patients that had elevated transaminases and the patients that did not (p-value = 0.2).

Higher level of care was statistically associated with risk of transaminase elevation (p-value < 0.001). Patients with normal AST and ALT values were more likely to be seen in the ED and subsequently discharged with 61% of the patients versus 32% of the patients that had elevated transaminases.

In this patient population, 39.8% had elevated transaminases and 10.5% were vaccinated against SARS-CoV2. Vaccinated children had lower AST: median 17.5 interquartile range (14.2, 28) vs IQR (23.0, 52) (Figure 1). Among vaccinated children, the prevalence of transaminase elevation was 21.4% and among unvaccinated children, the prevalence was 42.0%, but this difference is not statistically significant. However, AST levels in unvaccinated patients were significantly higher (34.0 vs 17.5, p <0.001). There was no significant difference in mean ALT level. (Table 1).

**Table 1:** Demographics and Biomarkers at Time of Presentation by Vaccination Status.

|  | No vaccine (n=119) | Vaccine (n=14) | P-value |
| --- | --- | --- | --- |
| Transaminemia | 50 (42.0%) | 3 (21.4%) | 0.2 |
| AST (highest), median, IQR | 34.0 (23.0, 52) | 17.5 (14.2, 28) | <0.001 |
| AST elevations |  |  | 0.2 |
| None [Normal AST ≤ 46] | 85 (71.4) | 14 (100) | |
| Mild [Abnormal, less than 2x ULN] | 29 (24.4) | 0 |  |
| Moderate [Between 2-5x ULN] | 4 (3.4) | 0 |  |
| Severe [More than 5x ULN] | 1 (0.8) | 0 |  |
| AST/ALT ratio, median, IQR | 1.90 (1.36, 2.54) | 1.40 (1.06, 1.85) | 0.02 |
| ALT, median, IQR | 17.0 (12.0, 22.5) | 14.5 (10.8, 18.5) | 0.2 |
| Random glucose, median, IQR | 97.0 (89.0, 107.0) | 96.0 (88.0, 121.5) | 0.9 |
| Albumin, median, IQR | 4.50 (4.1, 4.8) | 4.35 (4.1, 4.8) | 0.8 |
| Alkaline phosphatase, median, IQR | 208 (113, 258) | 117.5 (60.8, 220) | 0.1 |
| Platelets, median, IQR | 267 (207, 368) | 298.5 (235.8, 362.3) | 0.5 |
| Total bilirubin, median, IQR | 0.40 (0.2, 0.6) | 0.40 (0.2, 0.6) | 0.8 |
| Age (months), median (interquartile range) (range) | 108 (26, 180) (Range: 1-216) | 186 (156, 204) (Range: 84-223) | 0.001 |
| Age group |  |  | 0.007 |
| 0-4 years | 34 (28.6%) | 0 |  |
| 5-11 years | 40 (33.6%) | 3 (21.4%) |  |
| 12-19 years | 45 (37.8%) | 11 (78.6%) |  |
| Hospital |  |  | 0.03 |
| Downstate | 32 (26.9%) | 8 (57.1%) |  |
| Kings County | 87 (73.1%) | 6 (42.9%) |  |
| Date presenting |  |  | 0.003 |
| Mar.-Dec. 2020 | 29 (24.4%) | 0 |  |
| Jan.-June 2021 | 10 (8.4%) | 0 |  |
| June-Nov. 2021 | 23 (19.3%) | 0 |  |
| Nov. 2021-Mar. 2022 | 57 (47.9%) | 14 (100%) |  |
| Black race | 90 (78.3%) | 12 (85.7%) | 0.7 |
| Non-Hispanic ethnicity | 105 (91.3%) | 13 (92.9%) | 1.0 |
| Male sex | 63 (52.9%) | 9 (64.3%) | 0.4 |
| Preexisting conditions | 52 (43.7%) | 11 (78.6%) | 0.01 |
| Asthma | 16 (13.4) | 6 (42.9) | 0.01 |
| BMI, median, IQR | 19.2 (16.8-24.5) | 25.6 (19.0, 27.0) | 0.02 |
| BMI z-score, median (IQR) | 0.47 (-0.40, 1.67) | 1.21 (0.24, 1.81) | 0.2 |
| BMI z-score, mean (SD) | 0.41 (1.75) | 1.02 (1.25) | 0.1 |
| BMI class |  |  | 0.2 |
| 0 | 68 (62.4%) | 6 (42.9%) |  |
| 1 | 20 (18.3%) | 3 (21.4%) |  |
| 2 | 21 (19.3%) | 5 (35.7%) |  |
| Missing | 10 (8.4%) | 0 |  |
| Presenting with GI symptoms | 63 (52.9) | 10 (71.4) | 0.2 |
| Presenting with nausea | 31 (26.1) | 3 (21.4) | 1.0 |
| Presenting with diarrhea | 14 (11.8) | 2 (14.3) | 0.7 |
| Presenting with decreased intake | 18 (15.1) | 1 (7.1) | 0.7 |
| Presenting with abdominal pain | 22 (18.5) | 5 (35.7) | 0.2 |
| Presenting with other GI symptoms | 22 (18.5) | 4 (28.6) | 0.5 |
| Level of care |  |  | 0.3 |
| ED and discharged | 56 (47.1) | 10 (71.4) |  |
| Floor | 45 (37.8) | 3 (21.4) |  |
| PICU | 18 (15.1) | 1 (7.1) |  |

P-value for continuous variables was from Wilcoxon test due to skewed distributions. P-value from categorical variable was from chi-square test.

**Figure 1:**
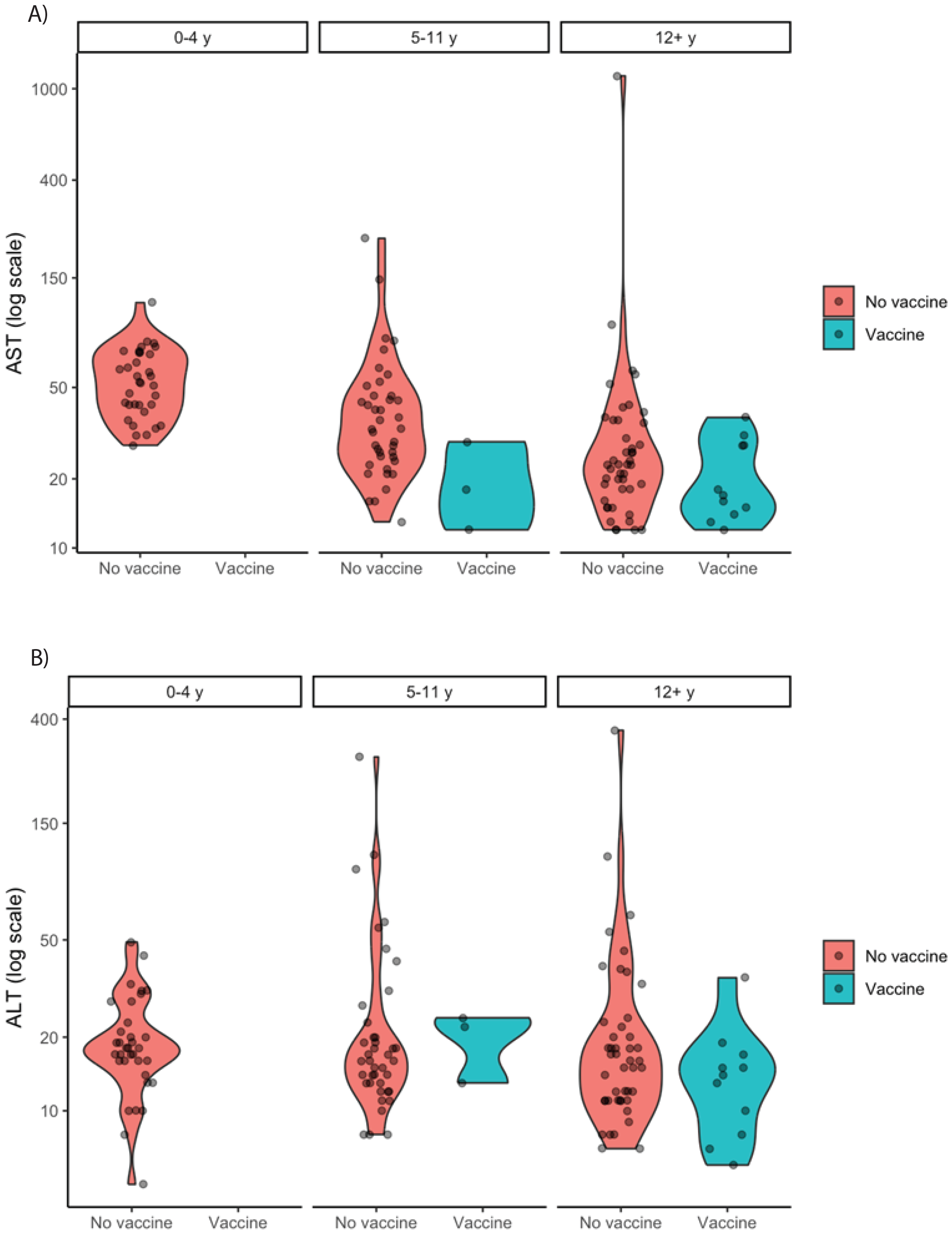
Violin Plot Summary of a) AST and b) ALT elevation by vaccination status.

Younger children were more likely to have elevated transaminases p 0.0001. Patients with normal transaminase levels were on average 5 years older and more likely to be 5-11 and 12+ years old. For each year of age, there were 7% fewer cases of transaminase elevation (PR = 0.93, 95% CI (0.89, 0.97)). Patients older than 12 years old had 62% lower risk of elevated transaminases than ages 0-11 years old. (Table 2) Rates of elevated transaminases appeared to increase by predominant variant type, with 9/29 (31%) of wild type window patients, 3/10 (30%) of alpha window patients, 9/23 (29.1%) delta window patients, and 32/71 (45.1%) for omicron window patients. AST/ALT ratio increased during the omicron window.

**Table 2:** Demographics and Results by Transaminase Elevation Status.

|  | No elevated transaminases (n=80) | Elevated transaminases (n=53) | P-value |
| --- | --- | --- | --- |
| Vaccinated | 11 (13.8%) | 3 (5.7%) | 0.1 |
| Antibody positive (vs. no testing or negative) | 23 (31.9) | 6 (12.2) | 0.01 |
| AST, median, IQR | 24 (18, 33)<br>Range 12-46 | 56 (42, 72)<br>Range (12-1137) | <0.001 |
| AST elevations |  |  |  |
| None [Normal AST ≤ 46] | 80 (100) | 19 (35.8) | |
| Mild [Abnormal, less than 2x ULN] | 0 | 29 (54.7) |  |
| Moderate [Between 2-5x ULN] | 0 | 4 (7.5) |  |
| Severe [More than 5x ULN] | 0 | 1 (1.9) |  |
| AST/ALT ratio, median, IQR | 1.9 (1.5, 2.3)<br>Range 0.6-8.40 | 2.1 (1.1, 3.2)<br>Range 0.32-6.31 | 0.8 |
| ALT, median, IQR | 13.5 (11.0, 17.0) | 27.0 (18.0, 41.0) | <0.001 |
| Glucose, median, IQR | 96.0 (89.0, 109.0) | 98.0 (88.0, 107.0) | 0.9 |
| Albumin, median, IQR | 4.55 (4.2, 4.8) | 4.50 (3.9, 4.8) | 0.6 |
| Alkaline Phosphatase, median, IQR | 181 (84.8, 241) | 225 (133, 269) | 0.05 |
| Platelets, median, IQR | 279 (217, 374) | 270 (197.5, 352.5) | 0.4 |
| Total bilirubin | 0.4 (0.2, 0.5) | 0.4 (0.2, 0.7) | 0.2 |
| Age (months), median (interquartile range) (range) | 144 (69, 201)<br>Range 1-216 | 60 (13, 144)<br>Range 1-223 | <0.001 |
| Age group |  |  | 0.002 |
| 0-4 years | 13 (16.2%) | 21 (39.6%) |  |
| 5-11 years | 25 (31.2%) | 18 (34.0%) |  |
| 12-19 years | 42 (52.5%) | 14 (26.4%) |  |
| Hospital |  |  | 0.98 |
| Downstate | 24 (30.0%) | 16 (30.2%) |  |
| Kings County | 56 (70.0%) | 37 (69.8%) |  |
| Date presenting |  |  | 0.55 |
| Mar.-Dec. 2020 | 20 (25.0%) | 9 (17.0%) |  |
| Jan.-June 2021 | 7 (8.8%) | 3 (5.7%) |  |
| June-Nov. 2021 | 14 (17.5%) | 9 (17.0%) |  |
| Nov. 2021-Mar. 2022 | 39 (48.8%) | 32 (60.4%) |  |
| Black race | 61 (79.2%) | 41 (78.8%) | 0.96 |
| Hispanic ethnicity | 6 (7.8%) | 5 (9.6%) | 0.76 |
| Male sex | 37 (46.2%) | 35 (66%) | 0.02 |
| Preexisting conditions | 38 (47.5%) | 25 (47.2%) | 0.97 |
| Asthma | 19 (23.8) | 3 (5.7) | 0.006 |
| BMI, median, IQR | 20.0 (17.7, 26.0) | 18.7 (16.5, 24.7) | 0.2 |
| BMI z-score, median (IQR) | 0.56 (-0.41, 1.79) | 0.79 (-0.22, 1.41) | 1 |
| BMI class |  |  | 0.3 |
| 0 | 44 (55.0%) | 30 (56.6%) |  |
| 1 | 12 (15.0%) | 11 (20.8%) |  |
| 2 | 19 (23.8%) | 7 (13.2%) |  |
| Missing | 5 (6.3%) | 5 (9.4%) |  |
| Presenting with GI symptoms | 48 (60.0) | 25 (47.2) | 0.2 |
| Presenting with nausea | 19 (23.8) | 15 (28.3) | 0.6 |
| Presenting with diarrhea | 10 (12.5) | 6 (11.3) | 0.8 |
| Presenting with diminished food intake | 15 (18.8) | 4 (7.5) | 0.07 |
| Presenting with other GI symptoms | 20 (25.0) | 6 (11.3) | 0.05 |
| Level of care |  |  | 0.001 |
| ED and discharged | 49 (61.3%) | 17 (32.1%) |  |
| Floor | 25 (31.2%) | 23 (43.4%) |  |
| PICU | 6 (7.5%) | 13 (24.5%) |  |

P-value for continuous variables was from Wilcoxon test due to skewed distributions. P-value from categorical variable was from chi-square test.

Children who were vaccinated presented during the Omicron-prevalent period (since November 2021), were more likely to have preexisting conditions (79% vs. 44%) more likely to be 5 years older at median (175 months vs. 102 months), and lower median AST/ALT ratio (1.4 vs. 1.9) (Table 1). Vaccinated children had a higher mean BMI than never-vaccinated children (25.6 vs 21.3) (Table 1).

In causal mediation analysis, 51% of the effect of the effect of the vaccine on transaminase elevation is mediated by a binary indicator for being age 12 or older, controlling for preexisting condition and presenting during omicron (p=0.04). In a second analysis, 69% of the effect of the effect of the vaccine on transaminase elevation is mediated by age in months, controlling for preexisting condition and presenting during omicron (p=0.02). However, the percentage of the effect of the effect of the vaccine on transaminase elevation is mediated by age in months controlling for preexisting condition and BMI class is not statistically significant (p=0.08).

## Discussion

While other studies have in part characterized the impact of SARS-CoV2 on the pediatric liver, this is the first reported work we are aware of to assess alterations to this behavior over variant windows, the first to assess vaccine protection, and the first to review this in a population of majority african descent and a correspondingly low risk of NAFLD. A focus on a minoritized and low-income population is also highly relevant, as this population remains most at risk of SARS-CoV2 infection^19^. We note: a significant age association (higher risk in younger children), a shift in characteristic of transaminase elevation over the variant windows, an absence of a BMI association in this primarily Black cohort, and a significant difference in AST elevation and likely difference in overall risk of transaminase elevation in vaccinated patients. Our most significant finding is the likely protective effect of vaccination, although due to the timing of our data and the strong age association of transaminase elevation risk, we were not able to determine significance in this study despite a wide practical variation and severity range between vaccinated and unvaccinated patients.

We have noted that pediatric patients with elevated transaminases were more likely to require higher level of care, although in the absence of a prospective study examining serial measurements and outcomes it is not clear if this reflects a risk factor or an outcome of severe illness. Interestingly, BMI, which has been felt to be a major risk factor both for SARS-CoV2 severity and transaminase elevation secondary to non-alcoholic steatohepatitis (NASH) or non-alcoholic fatty liver disease (NAFLD) ^20, 21^, was not relevant, displaying no increased risk of transaminase elevation. However, as our hospital center serves a patient population with atypical demographics (see high percentage of Black race in demographics) this may be an outlier reflecting a specific resistance to NASH/NAFLD due to polymorphisms such as PNPLA-3 or MBOAT7^22^.

Interestingly, children that were vaccinated presented with less incidence of elevated transaminases than non-vaccinated children (42 versus 21%, p-value 0.2%), although it was not statistically significant due to previously discussed limitation on the age range of vaccinated patients, and the strong correlation of young age and elevated transaminases. Level of transaminase elevation was significantly lower in vaccinated patients (AST 17.5 vs. 34.0, p = 0.002) suggesting a degree of protective impact. This is likely a function of age/limitations on when vaccines were available in young children, and it seems highly likely that in a larger study a strong vaccine protection would be noted.

Our study returned other notable findings. First, while we had theorized that GI infection may predispose to transaminase elevation, no connection was noted (Table 2). Second, we observed that with variant progression and population immunity levels there were shifts to the AST/ALT ratio (The DeRitis ratio) The ratio represents the time course and aggressiveness of disease that would be predicted from the relatively short half-life of AST (18 h) compared to ALT (36 h), and can be used to infer origin of transaminase elevation^23^. As the AST/ALT ratio increased over the time (Figure 2), it may be that transaminase elevations are increasingly of extra hepatic origin. Acute illness can also increase AST due to his extra hepatic presence in the muscle, heart and kidney, which aligns with our finding that level of care was linked to transaminase elevation.

**Figure 2:**
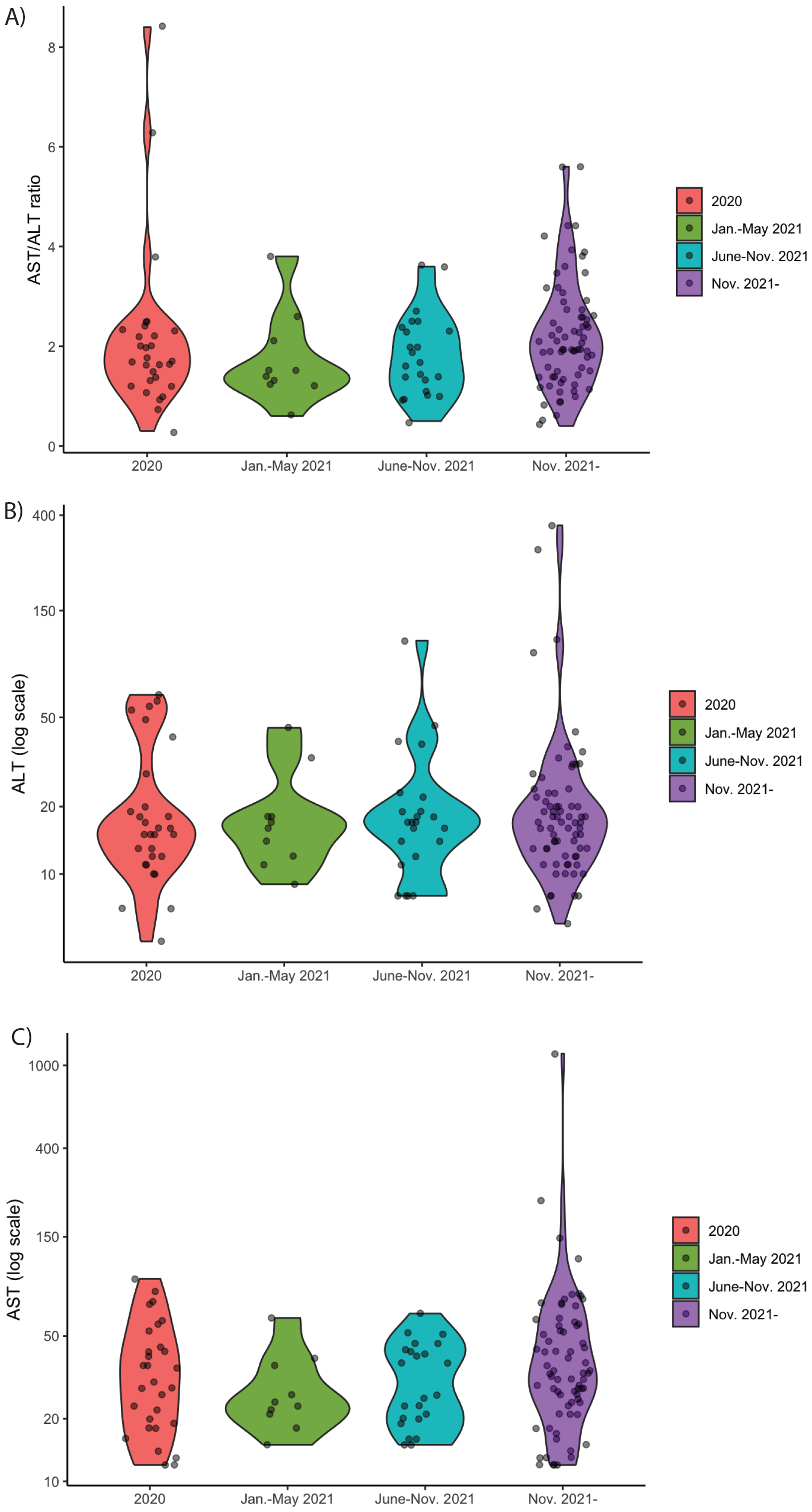
Change in Transaminase Elevation Behavior by Variant Window. A) AST/ALT ratio by window. B) ALT log scale by window. C) AST log scale by window.

In summary, this retrospective cross-sectional study of SARS-CoV2 associated hepatic injury and transaminase elevation has clearly identified major risk factors of young age and disease severity, and highly suggests a strong protective effect from vaccination. It also suggests that increasingly transaminase elevation in this population may not be primarily hepatic, although further study is needed. This study was limited by its retrospective nature, lack of reliably trended results, the bias towards severity inherent in requiring lab work (as frequently mild infections do not result in serum testing), and population size. With that said, several patients had transaminase elevations of significant size, suggesting risk in rare cases for fulminant hepatitis. This work presents a further argument towards the value of pediatric vaccination, although additional larger studies are indicated.

## Data Availability

Data available on request.

## Abbreviations

(AST): Aspartate Aminotransferase
(AST): Alanine Aminotransferase
(SES): Socioeconomic Status

